# Deducing the Dose-response Relation for Coronaviruses from COVID-19, SARS and MERS Meta-analysis Results

**DOI:** 10.1101/2020.06.26.20140624

**Authors:** Xiaole Zhang, Jing Wang

## Abstract

The fundamental dose-response relation is still missing for better evaluating and controlling the transmission risk of COVID-19. A recent study by Chu et al. has indicated that the anticipated probability of viral infection is about 12.8% within 1 m and about 2.6% at further distance through a systematic review and meta-analysis. This important information provides us a unique opportunity to assess the dose-response relation of the viruses, if reasonable exposure dose could be estimated. Here we developed a simple framework to integrate the *a priori* dose-response relation for SARS-CoV based on mice experiments, and the recent data on infection risk and viral shedding, to shed light on the dose-response relation for human. The developed dose-response relation is an exponential function with a constant *k* in the range of 6.19×10^4^ to 7.28×10^5^ virus copies. The result mean that the infection risk caused by one virus copy in viral shedding is about 1.5×10^−6^ to 1.6×10^−5^. The developed dose-response relation provides a tool to quantify the magnitude of the infection risk.

---

Control of the spread of Corona Virus Disease 2019 (COVID-19) is the urgent challenge throughout the world. It is considered that the causal pathogen, severe acute respiratory syndrome coronavirus 2 (SARS-CoV-2), could spread via the transmission of virus-laden respiratory particles shed from infected individuals ^1^. However the fundamental dose-response relation is still missing for better evaluating and controlling the transmission risk of the disease.

A recent study by Chu et al. ^2^ has indicated that the anticipated probability of viral infection is about 12.8% within 1 m and about 2.6% at further distance through a systematic review and meta-analysis on the betacoronaviruses causing severe acute respiratory syndrome (SARS), Middle East Respiratory Syndrome (MERS) and COVID-19. This important information provides us a unique opportunity to assess the dose-response relation of the viruses, if reasonable exposure dose could be estimated. Here we developed a simple framework to integrate the *a priori* dose-response relation for SARS-CoV ^3^ based on mice experiments, and the recent data on infection risk ^2^ and viral shedding ^4^, to shed light on the dose-response relation for human.

Watanabe et al. ^3^ found that the exponential model *p*=1−exp(−*d*/*k*) could well depict the dose-response relation based on the experiments challenging mice with recombinant SARS-CoV variants ^5^ and Murine Coronavirus Strain 1 (MHV-1) ^6^, where *p* is the infection probability, *d* is the exposure dose, and *k* is the pathogen dependent parameter. With a little mathematics, we could derive that *p* ∼ *d/k*, when the infection risk is below 15%, with the difference between *p* and *d*/*k* smaller than 8%. We assume that the exponential model remains applicable for the dose-response relation for human.

Recently, Leung et al. ^4^ investigated the respiratory shedding of coronaviruses (NL63, OC43, HKU1 and 229E) in exhaled breath in real-life situations with breathing and coughing. The results have shown that about 30% to 40% of the symptomatic individuals in the tests produced viral shedding (*E*_*virus*_) in respiratory particles, with about 10^2^ to 10^5^ virus copies in samples of 30 min without wearing masks, with a geometric mean of about 10^4^.

An overall effective dilution rate is required to convert the viral shedding into the exposure dose at various distances. The overall effective dilution rate should be a combination of various factors, e.g. the dilution when the exhaled air is mixed with the ambient air and the possibility for the exposed person to actually inhale the contaminated plume. The meta-analysis study ^2^ offered a plausible way to estimate this factor. It was shown that the chance of viral infection decreased by about a factor of 5 from within 1 m (12.8%) to further distance (2.6%). According to the exponential model, the dose-response relation is nearly linear within the range of *p* < 15% (*k* is a constant), which suggests that the viral shedding from an infected individual should also be effectively diluted by about the same magnitude. “Further distance” is likely within 2 or 3 meters considering the confined indoor space, so we use 5 times per meter as the effective dilution factor (*f*_*dilu*_) to estimate the exposure dose. It was reported in the meta-analysis ^2^ that the duration of exposure varied from any duration to a minimum of 1 h. Here we use 1 h as a representative duration (*t*_*expo*_), which is close to the total duration of close contact between a nurse / health worker and a patient per day ^7^. The exposure (virus copies) at further distance was estimated as *d* =*E*_*virus*_/(*f*_*dilu*_) *t*_*expo*_.

Monte Carlo simulations were conducted to estimate the exposure dose. Two different fractions (40%, 100%) of the infected individuals for positive viral shedding (*E*_*virus*_, virus copies per hour) and two distributions log_10_(*E*_*virus*_)∼*Normal*(4, 0.5) and log_10_(*E*_*virus*_)∼*Uniform*(3, 5), namely four different combinations of the configurations were utilized to estimate the exposure dose and the corresponding dose-response relation, and to evaluate the sensitivities of the estimations to the configurations. One million values of *E*_*virus*_ were generated for each setting. The deposition of the particles in the respiratory system was estimated based on the size distribution ^8^ of particles generated by breathing and coughing and the deposition model for bioaerosols^9^. The overall deposition ratio was about 90%. The dose-response relation was estimated based on 5 different contribution levels (0.1, 0.25, 0.5, 0.75 and 1) of airborne virus-laden particles to the total dose from both exposure to airborne viruses and contact transmission.

The final dose-response estimations for *k* are from 6.19×10^4^ to 7.28×10^5^ copies dependent on the contribution of the airborne virus-laden particles to the total dose as shown in appendix Figure 1 to 4. The variations among different configurations are limited, which suggests the stability of the estimations. The results mean that the infection risk caused by one virus copy in viral shedding is about 1.5×10^−6^ to 1.6×10^−5^. Watanabe et al. utilized the plaque-forming units (PFU) to quantify the dose. A previous study on SARS-CoV ^10^ showed that about 300 viral genome copies were present per PFU. Then *k* in the Watanabe model ^3^ was about 1.23×10^5^, close to our estimation with 50% contribution from airborne particles to the total dose. The results suggest that the experiments based on mice could provide reasonable insight for the human infection risk. However, the conversion factor from PFU to virus copies is still uncertain and needs further evaluation.

**Figure 1.**
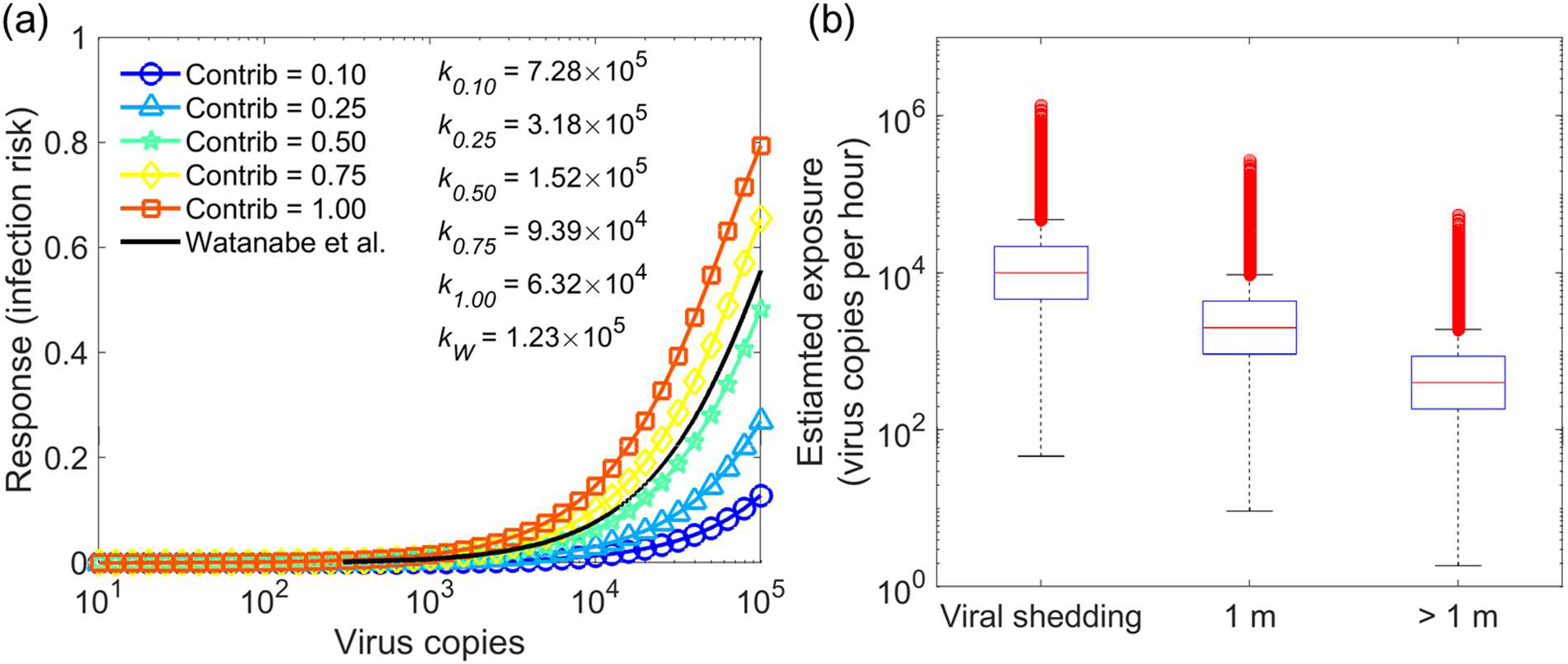
Log-normal distribution of viral shedding log_10_(*E*_*virus*_)∼*Normal*(4, 0.5), with 40% positive viral shedding. (a) The estimated dose-response relations based on the different contributions (0.1, 0.25, 0.5, 0.75 and 1) of the airborne virus-laden particles to the total dose from both exposure to airborne viruses and contact transmission; (b) viral shedding and exposure dose for 1 hour duration, zero values are not shown in the figure.

**Figure 2.**
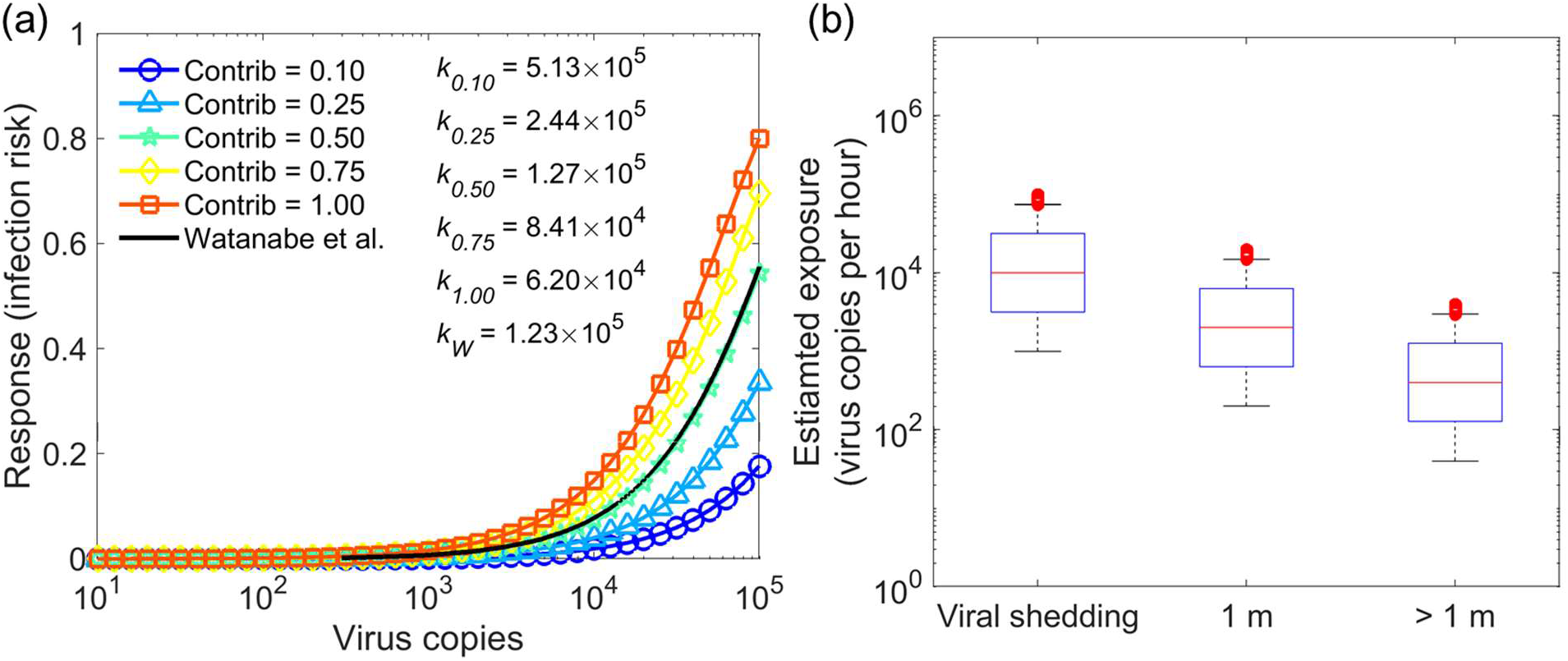
Log-uniform distribution of viral shedding log_10_(*E*_*virus*_)∼*Uniform*(3, 5), with 40% positive viral shedding. (a) The estimated dose-response relations based on the different contributions (0.1, 0.25, 0.5, 0.75 and 1) of the airborne virus-laden particles to the total dose from both exposure to airborne viruses and contact transmission; (b) viral shedding and exposure dose for 1 hour duration, zero values are not shown in the figure.

**Figure 3.**
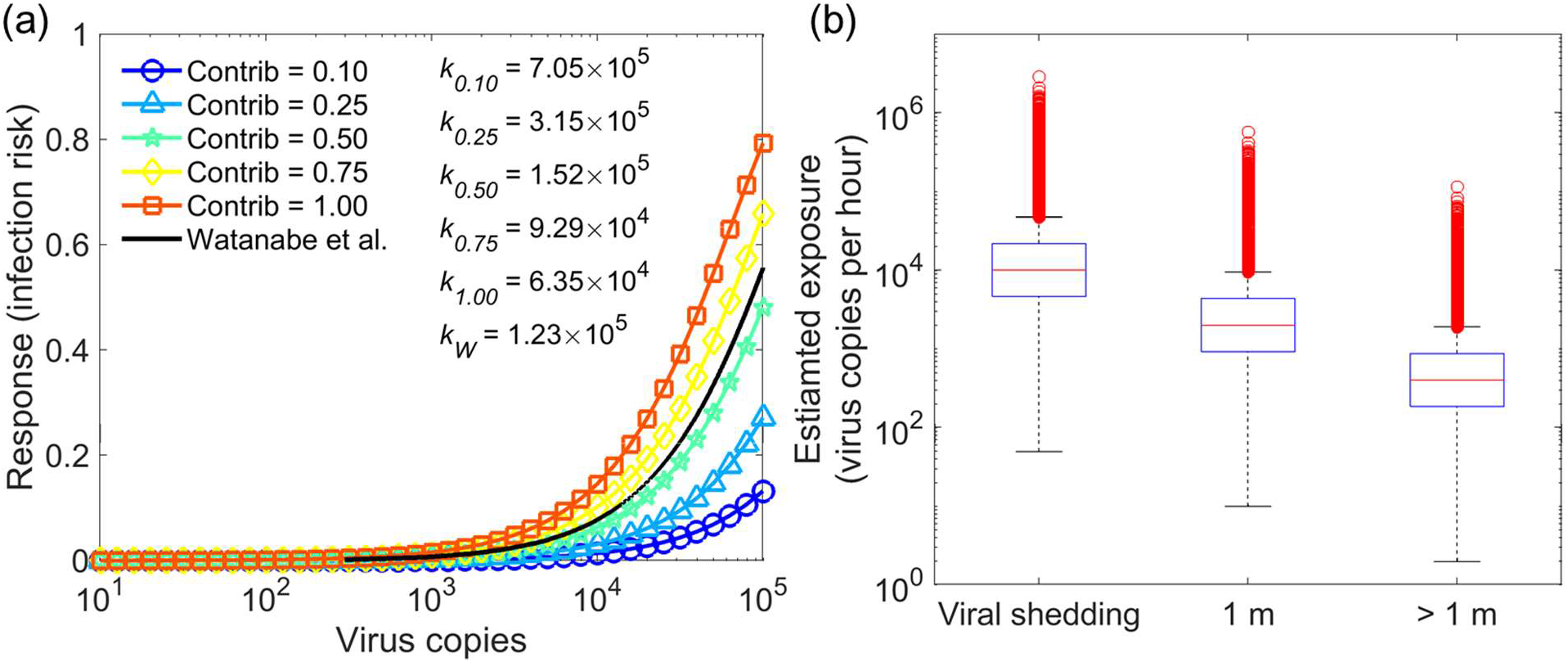
Log-normal distribution of viral shedding log_10_(*E*_*virus*_)∼*Normal*(4, 0.5), with 100% positive viral shedding. (a) The estimated dose-response relations based on the different contributions (0.1, 0.25, 0.5, 0.75 and 1) of the airborne virus-laden particles to the total dose from both exposure to airborne viruses and contact transmission; (b) viral shedding and exposure dose for 1 hour duration, zero values are not shown in the figure.

**Figure 4.**
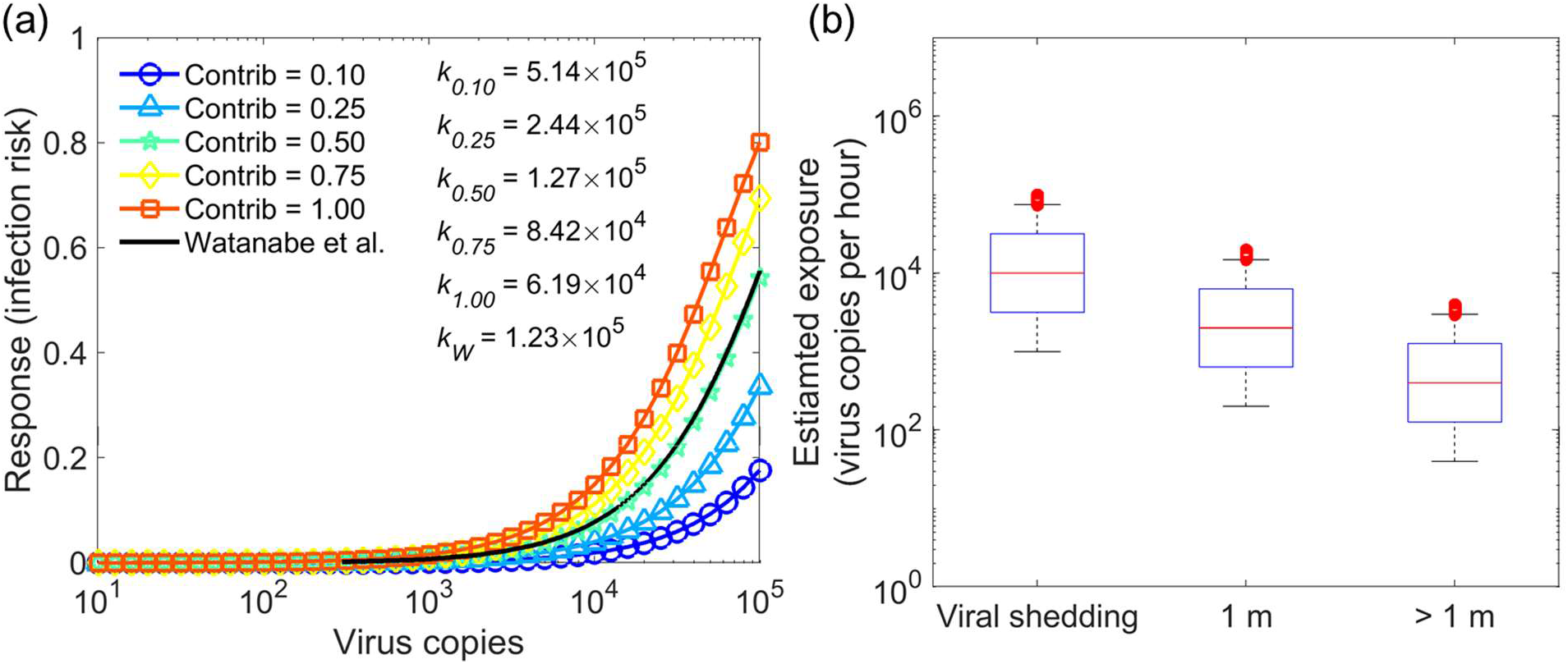
Log-uniform distribution of viral shedding log_10_(*E*_*virus*_)∼*Uniform*(3, 5), with 100% positive viral shedding. (a) The estimated dose-response relations based on the different contributions (0.1, 0.25, 0.5, 0.75 and 1) of the airborne virus-laden particles to the total dose from both exposure to airborne viruses and contact transmission; (b) viral shedding and exposure dose for 1 hour duration, zero values are not shown in the figure.

The framework is kept as simple as possible to avoid unnecessary uncertainties. The developed dose-response relation provides a tool to quantify the magnitude of the infection risk and can be used in model assessment of the infection risk in specific cases ^11^. The meta-analysis and evidence based study provide a new opportunity to estimate the human dose-response relation. In the future, the uncertainties could be reduced by using new technology, e.g. personal IoT (internet of things) devices, to record the distance, exposure duration, and environmental factors, which could greatly improve the quality of exposure dose estimation and thus the dose-response relation.

## Data Availability

All the data referred to in the manuscript are from published literature.

## Appendix

